# Association of immunoglobulin E levels with glioma risk and survival

**DOI:** 10.1101/2024.05.09.24307132

**Authors:** Geno Guerra, Taishi Nakase, Linda Kachuri, Lucie McCoy, Helen M. Hansen, Terri Rice, Joseph. L. Wiemels, John K. Wiencke, Annette M. Molinaro, Margaret Wrensch, Stephen S. Francis

## Abstract

**Background:** Previous epidemiologic studies have reported an association of serum immunoglobulin E (IgE) levels with reduced glioma risk, but the association between IgE and glioma prognosis has not been characterized. This study aimed to examine how sex, tumor subtype, and IgE class modulate the association of serum IgE levels with glioma risk and survival.

**Methods:** We conducted a case-control study using participants from the University of California, San Francisco Adult Glioma Study (1997-2010). Serum IgE levels for total, respiratory and food allergy were measured in adults diagnosed with glioma (n=1319) and cancer-free controls (n=1139) matched based on age, sex, and race and ethnicity. Logistic regression was adjusted for patient demographics to assess the association between IgE levels and glioma risk. Multivariable Cox regression adjusted for patient-specific and tumor-specific factors compared survival between the elevated and normal IgE groups. All statistical tests were 2-sided.

**Results:** Elevated total IgE was associated with reduced risk of IDH-wildtype (RR=0.78, 95% CI: 0.71-0.86) and IDH-mutant glioma (RR=0.73, 95% CI: 0.63-0.85). In multivariable Cox regression, positive respiratory IgE was associated with improved survival for IDH-wildtype glioma (RR=0.79, 95% CI: 0.67-0.93). The reduction in mortality risk was significant in females only (RR=0.75, 95% CI: 0.57-0.98) with an improvement in median survival of 6.9 months (P<.001).

**Conclusion:** Elevated serum IgE was associated with improved prognosis for IDH-wildtype glioma, with a more pronounced protective effect in females than males, which has implications for the future study of IgE-based immunotherapies for glioma.

## INTRODUCTION

Glioma is a primary malignant brain tumor that causes 11,000 cancer-related deaths in the United States each year^1,2^. IDH-wildtype glioblastoma (IDH-WT GBM), the most common subtype, carries a median overall survival of 14 months^3^. GBM has a clear sex bias, where males are more likely to develop glioma and have poorer outcomes than females^4^. In the last two decades, epidemiologic studies have highlighted a putative role of immune-related factors, including viral infections^5,6^, allergic conditions^7^ and autoimmune disorders^8^ in gliomagenesis.

A history of allergies is generally associated with a 20% to 40% decrease in the risk of developing glioma^9–12^. Using serum immunoglobulin E (IgE) concentrations as a biomarker for allergic sensitization, recent studies have reported an inverse association between elevated IgE levels and glioma risk, where banked serum studies show that this association exists up to 20 years prior to cancer diagnosis^10,11,13–15^. The association is consistently more robust among females, especially for respiratory IgE and GBM^13,15^. In a small cohort of GBM patients, elevated IgE levels have previously been associated with improved survival^16^, suggesting that IgE-mediated immune responses may not only suppress tumor development but also slow cancer progression in the tumor microenvironment. Based on these observations, we hypothesized that the relationship between IgE levels and survival may be modified by sex, tumor subtype and IgE class.

In this study, we aimed to refine the relationship between IgE levels and glioma risk according to contemporary molecular classifications and analyze the impact of sex, tumor subtype and IgE class on the association between IgE levels and survival in adult glioma patients.

## METHODS

### Ethics

Patient samples and clinicopathological information were collected with informed consent and relevant ethical review board approval per the tenets of the Declaration of Helsinki from the Institutional Review Board of the University of California, San Francisco (UCSF), Human Research Protection Program in the Office of Ethics and Compliance (USA).

### Study Population

Collection of samples along with demographic and clinical information for participants of the UCSF Adult Glioma Study (AGS) has been previously described^7,11,17^. Briefly, adults 18 years or older with a newly diagnosed glioma between 1997-1999 (series 2), 2001-2004 (series 3), or 2006-2010 (series 4) who resided in the SF Bay Area were eligible for enrollment in the AGS study. Series 3 and 4 also included eligible adults seeking care at the UCSF Neuro-oncology clinic, regardless of primary residence. The diagnosis of glioma (ICDO-3 codes 9380-9480 or equivalent) was established through histology in all cases according to 2016 World Health Organization (WHO) guidelines. Controls recruited from the SF Bay Area were identified using random digit dialing and were frequency matched to cases based on age, sex, and self-reported race and ethnicity. Treatment information (including use of dexamethasone, temozolomide or other chemotherapy, and radiation) at time of, or prior to, blood draw was recorded for all cases. Follow-up is ongoing for surviving participants.

### IgE Measurements

IgE antibody measurements for total, respiratory and food allergy were measured using serum from a single post-diagnosis time point for each case or at the time of the enrollment interview for each control. The full details of IgE measurements have been described in previous analyses of AGS participants^10,11^. Briefly, IgE levels were measured using the Pharmacia Diagnostics UniCAP fluorescent “sandwich” assay^18^. Total IgE measured the sum of all IgE antibodies found in the blood. Total IgE levels <25 kilounits/liter were considered clinically “normal”, and levels ≥25 were considered “borderline” or “elevated” (henceforth termed “above normal”). The respiratory IgE panel (Phadiotop) was composed of 15 allergens that encompass 97% of respiratory atopic allergy. The food IgE panel (fx5) consisted of allergy tests for shellfish, peanut, milk, egg, tree nut, and codfish, which comprise most food allergies. Food or respiratory IgE levels <0.35 kilounit/liter were termed “negative”, and levels ≥0.35 were called “positive”.

### Statistical Analysis

#### Risk Analysis

Risk ratios (RR) for increased IgE levels in cases versus controls were estimated using parametric g-computation^19^ under a logistic regression framework. We applied an inverse probability of treatment weighted (IPTW) approach^20^ to balance baseline covariates between IgE groups. The propensity models for IgE status (above normal/normal or positive/negative) used logistic regression with age (continuous), sex (M/F) and self-reported race and ethnicity (white/other) as covariates. Balance between the IgE groups was assessed with standardized differences of the baseline covariates, using a threshold of 0.1 to indicate imbalance. The outcome model for glioma used weighted logistic regression with IgE status as the independent variable and AGS recruitment window (series 2, 3 or 4) as a categorical variable. Heterogeneity in sex-specific IgE effects was assessed using Cochran’s Q test and the I^2^ index.

#### Survival Analysis

Mortality risk for positive/above normal IgE cases versus negative/normal IgE cases was estimated with IPTW Cox proportional hazards regression^20^. The propensity scores for IgE status were estimated using logistic regression with age (continuous), sex (M/F), self-reported race and ethnicity (white/other) and chemotherapy use (yes/no) as balancing covariates. The IPTW Cox model for the estimation of mortality risk between IgE groups included adjustment for categorical covariates for dexamethasone use at blood draw, radiation use, type of surgery (resection/biopsy), 1p/19q co-deletion, *TERT* mutation, tumor grade (2, 3 or 4), and AGS recruitment window (series 2, 3 or 4). Heterogeneity in sex-specific IgE effects was assessed using Cochran’s Q test and the I^2^ index. Substantial heterogeneity was defined as an I^2^>50% or a Q test p value<0.05^21^. Kaplan-Meier analyses were used to obtain unadjusted percent survival distributions and generate survival curves for visualization. Follow-up time was calculated from the date of pathological diagnosis to either the date of death or the date of last known contact for censored patients.

Population attributable fraction (PAF) estimates among IDH-WT glioma cases for age at diagnosis (<58 years/≥58 years), surgery (resection/biopsy), chemotherapy use (yes/no), and IgE (above normal/normal) were derived from Cox proportional hazard regression models adjusted for sex (combined only), self-reported race and ethnicity (white/other), dexamethasone use at blood draw, radiation use, 1p19q codeletion, *TERT* mutation, and strata for tumor grade (2, 3 or 4). PAF estimates are reported for 6 months and 12 months of follow-up time.

All statistical analyses were conducted using R v4.0.3. Statistical significance was determined using P<0.05 as the threshold in all comparisons; all P values were 2-sided.

## RESULTS

### Characteristics of Study Participants

The analysis group consisted of 1319 glioma cases (928 *IDH* wildtype, 391 *IDH* mutant) and 1139 non-glioma controls with at least one IgE antibody measurement. Basic information about glioma cases stratified by *IDH* mutation status as well as non-glioma controls is summarized in **Table 1**. The prevalence of above normal total IgE levels was higher in males than in females for cases (53.5% vs. 40.0%; **Supplementary Table 1**) and controls (63.9% vs. 51.0%; **Supplementary Table 2**). Individuals with positive respiratory IgE were younger and more likely male than those testing negative. The cohort of glioma cases with dexamethasone use at blood draw had a similar proportion of above normal total IgE measurements compared to cases without dexamethasone use (46.4% vs. 49.0%, P=0.31). The proportion of individuals with above normal total IgE or positive allergen-specific IgE was comparable across the three recruitment series (**Supplementary Table 3**).

**Table 1:** Description of Participants: University of California, San Francisco Adult Glioma Study (1997-2010).

|  | IDH-wildtype<br>N (%) | IDH-mutant<br>N (%) | Controls<br>N (%) |
| --- | --- | --- | --- |
| Total | 928 (100) | 391 (100) | 1139 (100) |
| Median age (IQR) | 56 (18) | 40 (16) | 57 (23) |
| Sex |  |  |  |
| Male | 569 (61.3) | 230 (58.8) | 588 (51.6) |
| Female | 359 (38.7) | 161 (41.2) | 551 (48.4) |
| Race and ethnicity |  |  |  |
| Asian | 50 (5.4) | 28 (7.2) | 82 (7.2) |
| Black or African American | 24 (2.6) | 9 (2.3) | 43 (3.8) |
| White | 827 (89.1) | 342 (87.5) | 981 (86.1) |
| Other<sup>a</sup> | 27 (2.9) | 12 (3.1) | 33 (2.9) |
| UCSF AGS series |  |  |  |
| Series 2 ('97-'99) | 140 (15.1) | 60 (15.4) | 289 (25.4) |
| Series 3 ('01-'04) | 467 (50.3) | 208 (53.2) | 470 (41.3) |
| Series 4 ('06-'10) | 321 (34.6) | 123 (31.5) | 380 (33.4) |
| Total IgE<sup>b</sup> |  |  |  |
| Normal | 479 (51.6) | 204 (52.2) | 482 (42.3) |
| Borderline | 289 (31.1) | 135 (34.5) | 396 (34.8) |
| Elevated | 159 (17.1) | 51 (13.0) | 261 (22.9) |
| Respiratory IgE<sup>b</sup> |  |  |  |
| Negative | 586 (63.2) | 223 (57.0) | 663 (58.2) |
| Positive | 340 (36.6) | 167 (42.7) | 476 (41.8) |
| Food IgE<sup>b</sup> |  |  |  |
| Negative | 873 (94.1) | 378 (96.7) | 1049 (92.1) |
| Positive | 53 (5.7) | 12 (3.1) | 90 (7.9) |
| IDH<sup>p</sup> |  |  |  |
| Mutated | 0 (0) | 391 (100) | — |
| Wildtype | 928 (100) | 0 (0) | — |
| 1p/19q<sup>b</sup> |  |  |  |
| Codeleted | 9 (1.0) | 116 (29.7) | — |
| Non-codeleted | 835 (90.0) | 242 (61.9) | — |
| Grade<sup>b</sup> |  |  |  |
| II | 39 (4.2) | 181 (46.3) | — |
| III | 82 (8.8) | 135 (34.5) | — |
| IV | 728 (78.5) | 62 (15.9) | — |
| Days to blood draw (IQR) | 90 (93) | 99 (116) | — |
| Chemotherapy |  |  |  |
| Yes | 708 (76.3) | 253 (64.7) | — |
| No | 220 (23.7) | 138 (35.3) | — |
| Temozolomide |  |  |  |
| Yes | 626 (67.5) | 212 (54.2) | — |
| No | 302 (32.5) | 179 (45.8) | — |
| Radiation |  |  |  |
| Yes | 841 (90.6) | 241 (61.6) | — |
| No | 87 (9.4) | 150 (38.4) | — |
| Dexamethasone |  |  |  |
| Yes | 410 (44.2) | 89 (22.8) | — |
| No | 518 (55.8) | 302 (77.2) | — |

For quantitative variables, interquartile range (IQR) is reported. <sup>a</sup>Other includes Native Americans, Pacific Islanders and individuals with no reported race and ethnicity. <sup>b</sup>Sum may not equal the total due to individuals with missing values. Tumor-specific and treatment-specific variables are not applicable to controls.

### Serum IgE and IDH-specific glioma risk

We examined the association between serum IgE levels and glioma risk stratifying by sex and *IDH* mutation status (**Table 2**). Above normal total IgE was associated with a 22% lower risk of developing IDH-WT glioma (RR=0.78, 95% Confidence Interval (CI): 0.71-0.86, P<.001). Similar results were observed when restricting to males (RR=0.80, 95% CI: 0.71-0.89, P<.001) and females (RR=0.75, 95% CI: 0.64-0.89, P<.001). For IDH-mutant (IDH-MT) gliomas, an inverse association was also identified in the pooled analysis (RR=0.73, 95% CI: 0.63-0.85, P<.001), which remained in both males (RR=0.77, 95% CI: 0.64-0.92, P=0.005) and females (RR=0.68, 95% CI: 0.52-0.87, P=0.003). Similar results were obtained when restricting to grade IV IDH-wildtype gliomas (**Supplementary Table 4**).

**Table 2:**
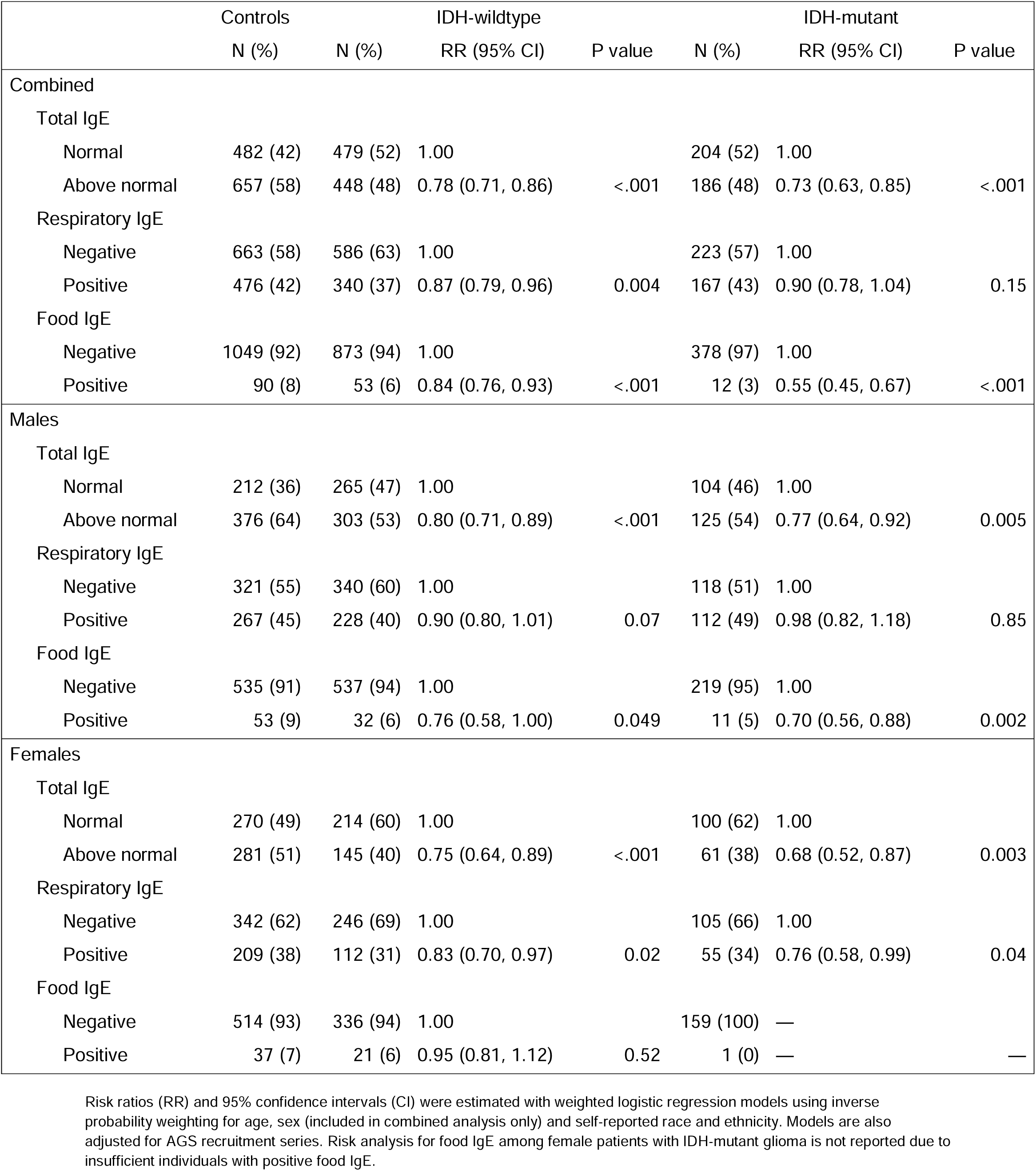
Associations between IgE levels and glioma risk stratified by sex and molecular subtype.

We found that testing positive for respiratory IgE was associated with a significant decrease in the risk of IDH-WT glioma (RR=0.87, 95% CI: 0.79-0.96, P=0.004), especially in females (RR=0.83, 95% CI: 0.70-0.97, P=0.02). This female-specific attenuation in risk was also observed for IDH-MT glioma (RR=0.76, 95% CI: 0.58-0.99, P=0.04), which showed evidence of sex-based heterogeneity in respiratory IgE effects (I^2^=60.2%; **Supplementary Table 5**). Positive food IgE was associated with a decrease in the risk of both IDH-WT (RR=0.84, 95% CI: 0.76-0.93, P<.001) and IDH-MT glioma (RR=0.55, 95% CI: 0.45-0.67, P<.001). This reduction in risk was restricted to males (IDH-WT: RR=0.76, P=0.049; IDH-MT: RR=0.70, P=0.002), with an estimated sex-based heterogeneity of I^2^=73.4% for IDH-WT glioma (**Supplementary Table 5**). Sensitivity analyses restricted to self-reported White participants yielded similar results to the pooled analyses (**Supplementary Table 6**).

### Serum IgE and overall survival

Among patients with IDH-WT gliomas, above normal total IgE was associated with improved survival (RR=0.84, 95% CI: 0.71-0.98, P=0.03; **Figure 1**, **Supplementary Table 7**). IDH-WT glioma cases with above normal total IgE levels showed a median 1.5 months longer survival than cases with normal levels (P=0.007). For cases with IDH-MT glioma, above normal total IgE levels were not associated with altered survival (RR=0.89, 95% CI: 0.63-1.26, P=0.50). The association between IgE and overall survival among patients with IDH-WT glioma was particularly pronounced for respiratory IgE (RR=0.79, 95% CI: 0.67-0.93, P=0.005), with positive cases demonstrating an increase in median survival of 2.2 months (P<.001). We did not observe a significant relationship between positive respiratory IgE and mortality risk among IDH-MT glioma patients (RR=0.92, 95% CI: 0.66-1.29, P=0.63). Food IgE was not associated with prognosis in either tumor subtype.

**Figure 1:**
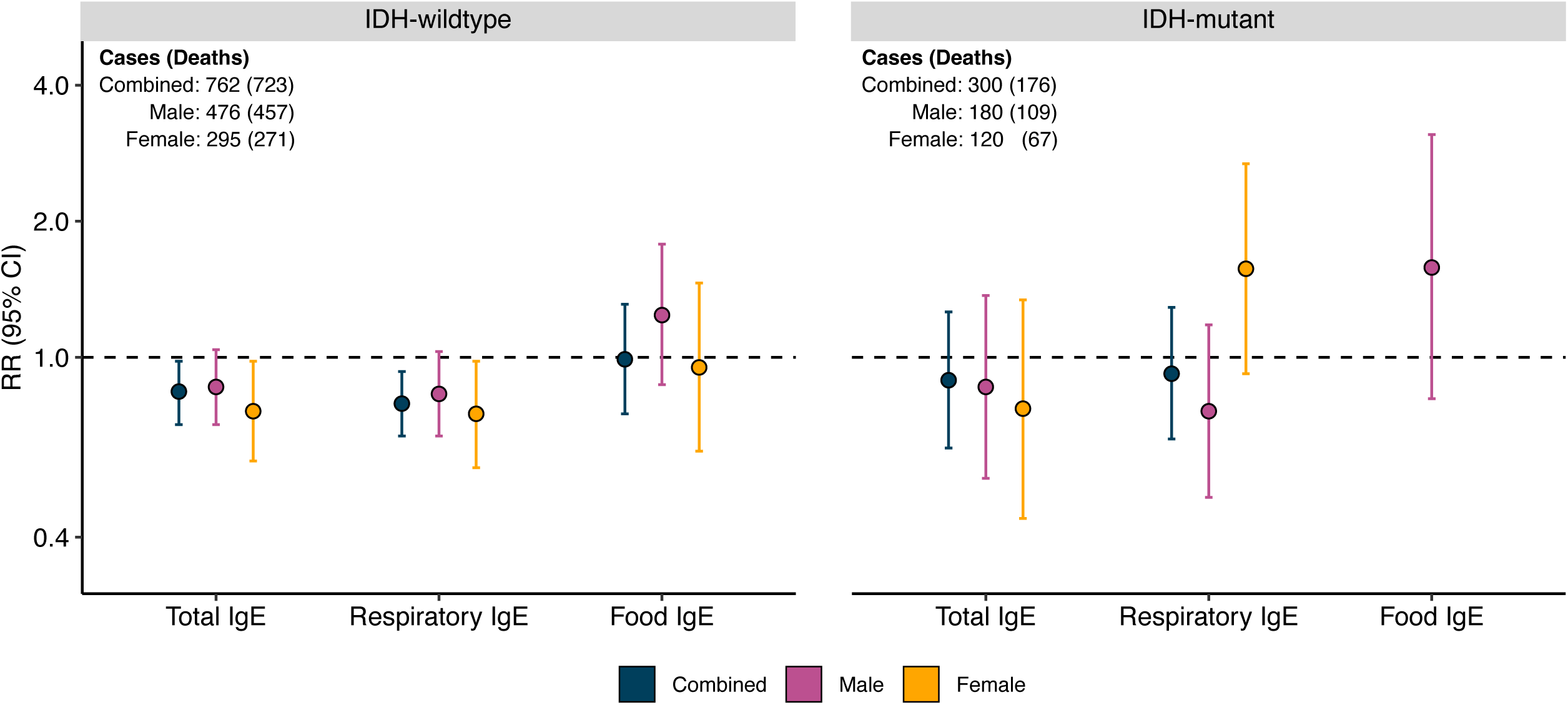
Associations between IgE levels and survival stratified by sex and molecular subtype. Risk ratios (RR) and corresponding 95% confidence intervals (CI) were estimated using IPTW Cox regression models. Propensity models include age, sex (included in combined analysis only), self-reported race and ethnicity, and chemotherapy use. Weighted Cox models were also adjusted for dexamethasone use, surgery type (resection/biopsy only), 1p19q codeletion status, *TERT* mutation, tumor grade and AGS recruitment series. The RR estimate for food IgE and IDH-mutant glioma (combined and female only) are not shown due to insufficient individuals with positive food IgE.

### Influence of sex on survival outcomes

Next, we assessed whether survival associations varied by sex (**Figure 2**, **Supplementary Table 7**). For IDH-WT glioma, above normal total IgE was associated with improved prognosis in females (RR=0.76, 95% CI: 0.59-0.98, P=0.04), but not in males (RR=0.86, 95% CI: 0.71-1.04, P=0.11). In females, median survival in patients with above normal total IgE was 6.2 months longer than in patients with normal levels (P=0.009). In males, the difference in median survival between normal and above normal total IgE groups was 1.2 months (P=0.04). Results were similar when restricting to grade IV IDH-WT gliomas (**Supplementary Table 8**), with above normal total IgE associated with a significantly lower mortality risk among females (RR=0.73, 95% CI: 0.55-0.97, P=0.03), but not among males (RR=0.86, 95% CI: 0.71-1.05, P=0.14).

**Figure 2:**
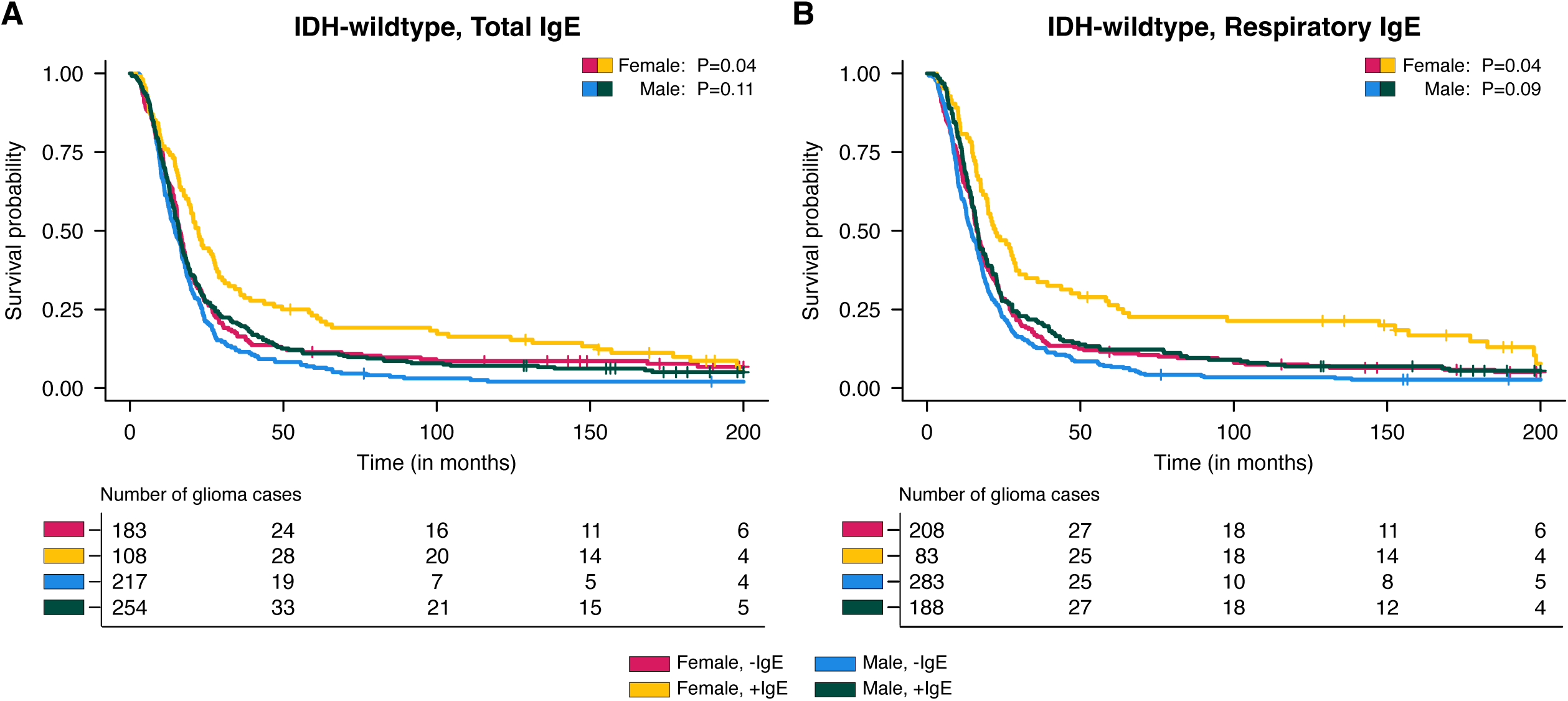
Kaplan-Meier curves for adult glioma cases with serum IgE concentrations. Percent survival distributions for adults with glioma stratified by sex and categorical serum IgE concentration for (**A**) IDH-wildtype glioma and total IgE and (**B**) IDH-wildtype glioma and respiratory IgE. +IgE = above normal total IgE or positive respiratory IgE.-IgE = normal total IgE or negative respiratory IgE. P values are shown for the sex-stratified Kaplan-Meier curves.

For IDH-WT glioma, females who were positive for respiratory IgE had a 25% decrease in overall mortality risk (RR=0.75, 95% CI: 0.57-0.98, P=0.04) and a median 6.9 months longer survival than those with negative antibody tests (P<.001). The magnitude of the effect was smaller and not statistically significant for males (RR=0.83, 95% CI: 0.67-1.03, P=0.09), with the improvement in median survival limited to 2.0 months (P=0.01). Evidence of sex-based heterogeneity in overall survival was observed for IDH-MT glioma and respiratory IgE (I^2^=75.8%, **Supplementary Table 9**), although the association between positive respiratory IgE and mortality did not achieve statistical significance in either females (RR=1.57, 95% CI: 0.92-2.68, P=0.10) or males (RR=0.76, 95% CI: 0.49-1.18, P=0.22). Although attenuated, similar results were obtained when restricting to self-reported White patients (**Supplementary Table 10**).

### Quantification of population-level impact on IDH-WT survival

Among patients with IDH-WT gliomas, the PAF at 6 months follow-up for positive respiratory IgE (PAF_rIgE_=0.14, P=0.004) exceeded that for both chemotherapy (PAF_chemo_=0.08, P=0.02) and surgery (PAF_surg_=0.08, P=0.003), but was less than that for age at diagnosis (PAF_age_=0.25, P<.001) (**Figure 3**, **Supplementary Table 11**). In contrast, the contribution of total IgE was more similar to treatment factors (PAF_tIgE_=0.08, P=0.04; **Supplementary** Figure 1). In females, elevated total IgE (PAF_tIgE_=0.15, P=0.03) and respiratory IgE (PAF_rIgE_=0.19, P=0.01) had a substantially larger impact on mortality risk than treatment factors, whereas in males, IgE did not significantly contribute to overall survival (PAF_tIgE_=0.06, P=0.17; PAF_rIgE_=0.11, P=0.08). At 12 months follow-up, PAF estimates for IgE were generally attenuated, but remained statistically significant for females (**Supplementary Table 11**).

**Figure 3:**
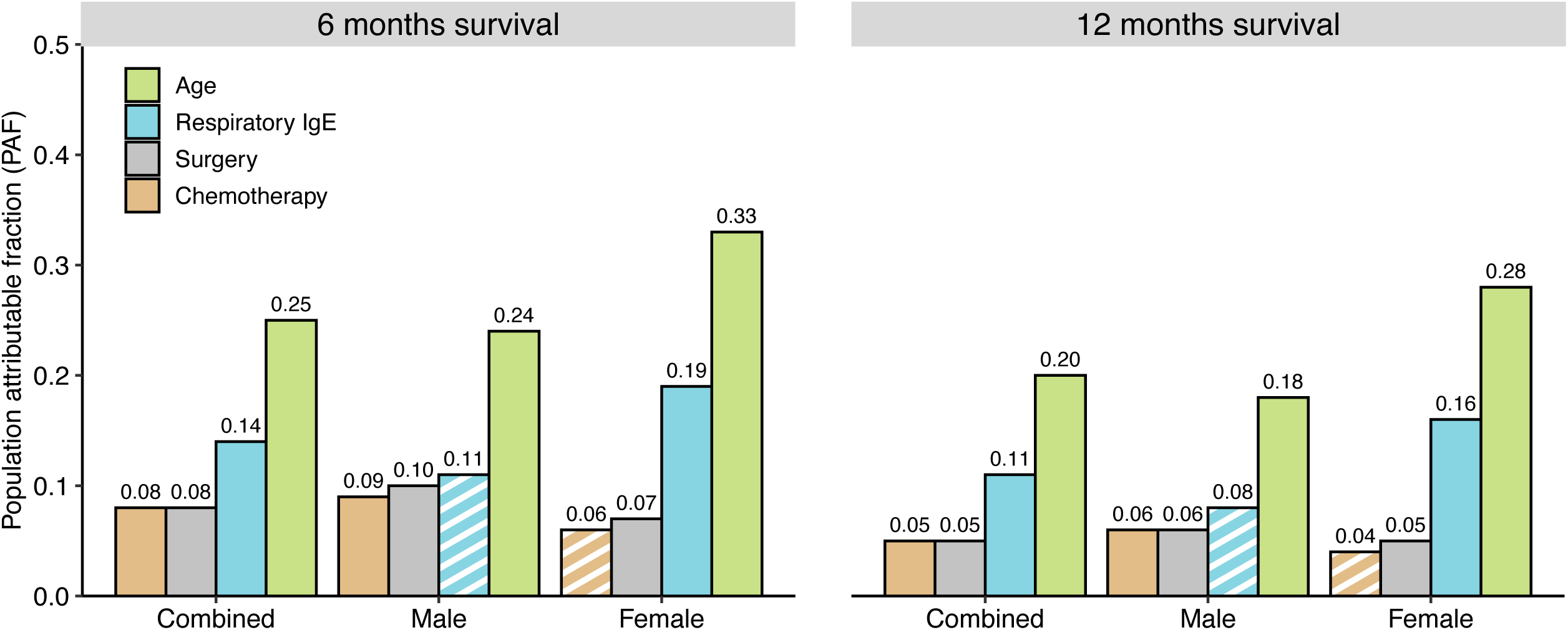
Population attributable fractions (PAF) for IDH-WT survival stratified by sex and follow-up time. PAF estimates for age at diagnosis (<58 years/≥58 years), surgery (resection/biopsy), chemotherapy use (yes/no), and respiratory IgE (positive/negative) were derived from Cox proportional hazard regression models adjusted for sex (combined only), self-reported race and ethnicity, radiation use, 1p19q codeletion status, *TERT* mutation status, and strata for tumor grade. PAF estimates are reported for 6 months and 12 months of follow-up time. Estimates that do not achieve P<0.05 are indicated by a striped pattern.

## DISCUSSION

We found that elevated levels of total IgE were associated with reduced risk of both IDH-WT and IDH-MT glioma, recapitulating the results of several previous studies^10,11,13,14^. In survival analyses, we demonstrated that elevated post-diagnosis serum IgE levels were associated with improved outcomes among patients with IDH-WT tumors. We identified a strong protective effect in IDH-WT female patients, where respiratory IgE seropositivity was associated with a 25% reduction in mortality risk resulting in approximately 7 months of additional survival time. This substantial improvement in median survival rivals the benefit of the principal glioma therapeutic, temozolomide^22^. Similarly, we observed a higher PAF for positive respiratory IgE than either chemotherapy or surgery in females with IDH-WT glioma. Together, these results highlight substantial differences in immune factors relating to both subtype-specific and sex-specific glioma prognosis.

The mechanism underlying the link between IgE-mediated atopic disease and glioma susceptibility remains unclear. The current prevailing hypothesis is that atopy induces a state of enhanced natural immunosurveillance, which promotes the detection and removal of malignant cells^23,24^. Though best known for its roles in allergy and protection against parasitic infections, IgE has been shown to exhibit both direct and cell-mediated effects against tumor cells^24^. These effects include the reprogramming of immunosuppressive tumor-associated macrophages and the activation of antibody-mediated tumor cell cytotoxicity and phagocytosis. For instance, in murine tumor models, the administration of IgE antibodies has been shown to recruit and re-educate immunosuppressive tumor-associated macrophages toward a tumoricidal phenotype via a TNFα/MCP-1 cascade^25,26^. The association between elevated IgE and improved IDH-WT prognosis suggests that some of these IgE-mediated immune responses may have direct antitumor activity within the central nervous system (CNS).

Recent research has shown that allergic airway inflammation can induce a pro-inflammatory-like state in microglia that reduced tumor growth in mice models^27,28^. Additionally, previous studies have shown that patients with hyper IgE syndrome (HIES) commonly develop white matter hyperintensities in the cerebral hemispheres^29^, suggesting that IgE-mediated autoimmunity is active in the CNS. Intriguingly, some forms of HIES have been shown to arise from dominant negative mutations in the signaling transduction and activator of transcription 3 (STAT3) gene^30^, which has been linked to many cancers including glioma^31,32^. While the role of *STAT3* and the JAK/STAT pathway has been well studied in IDH-WT GBMs^33–35^, its significance in IDH-MT glioma is less clear. Further research is needed to investigate the etiologic role of IgE in gliomagenesis, its interaction with genomic alterations and its potential as a therapeutic intervention.

While sex-specific differences in respiratory IgE and GBM risk have been previously reported^13,15^, this study is the first to suggest sexual dimorphism in the link between IgE and GBM survival. It is well established that males and females exhibit distinct immunological responses to foreign and self-antigens, which contribute to differences in susceptibility to autoimmune diseases, malignancies and allergies^36^. While adolescent males tend to be more affected by allergies and have higher allergen-specific IgE sensitization than females, rates of atopy and allergic disease in females increase during puberty to levels comparable to that in males^37–39^. Although the mechanisms by which sex hormones modulate allergy-induced inflammation are complex, experimental studies suggest that estrogen enhances immunological recall responses while testosterone dampens such responses^36,40^. Estrogen is thought to promote a Th2 cell bias^36,41^, which may enhance allergic inflammation and promote direct anti-GBM activity via IgE antibody dependent cellular cytotoxicity (ADCC)^42^. Intriguingly, in female patients with IDH-MT glioma, positive respiratory IgE appeared to increase mortality risk, although the association did not achieve statistical significance. This finding should be interpreted with caution and further replicated, but it suggests the possibility of female sex hormones interacting with tumor-specific immune features to modify the association between IgE and overall survival. Functional studies are needed to elucidate possible mechanisms by which the differential expression of sex hormones between males and females influences IgE-related immune responses against glioma.

Although we observed improved survival among IDH-WT patients with positive respiratory IgE, elevated food IgE was not associated with mortality risk. Isolated associations between respiratory IgE levels and glioma risk were observed in two previous studies^13,15^, suggesting that the effect of respiratory IgE may reflect the mechanism by which allergic inflammation influence glioma development and progression. It may be that specific respiratory allergens share epitopes with tumor antigens, such that adaptive immune cells primed by respiratory allergens recognize glioma cells more efficiently and have stronger antitumor effects. However, this hypothesis fails to account for previous reports of an inverse association between contact allergy, a delayed-type hypersensitivity reaction of the skin, and brain cancer^9,43^, which broadly indicate that the antitumor activity of allergic inflammation may not be linked to specific allergens or IgE itself. Furthermore, we observed a reduction in glioma risk in patients with either positive respiratory IgE or positive food IgE, suggesting that IgE-related effects on gliomagenesis are potentially independent of the class of IgE antibodies. Elevated food IgE was less common than respiratory IgE (6% vs. 41%); therefore, our study may lack statistical power to detect an association with mortality.

In addition to small sample sizes in some stratified analyses, our study has several limitations. Our cohort was initially categorized using WHO 2016 criteria and lacks some somatic mutations necessary for WHO 2021 classification. Nevertheless, *IDH* mutation captures the most prognostically significant glioma feature. Blood samples used to measure IgE antibody levels were collected a median of 95 days post-surgery among cases and may be influenced by the immunosuppressive effects of glioma treatment or the tumor itself. While there is some potential for this to confound the protective effect of IgE on glioma susceptibility, plasma IgE levels collected as much as 20 years before diagnosis have been reported to be associated with glioma risk^13,14^. Furthermore, in our analyses, adjustment for dexamethasone use did not substantively alter the effect of IgE levels on survival. Although use of dexamethasone and temozolomide were well documented in our cohort, we did not have information on the use of other immunomodulatory medications that have been associated with glioma risk among individuals with a history of allergies.

In this study we replicated the established association between elevated serum IgE and reduced glioma risk in the largest cohort to date. We also discovered that elevated IgE is associated with improved survival, especially in females, where the population attributable fraction to survival time was greater than the effects of treatment. These analyses were rigorously adjusted for potential imbalance in baseline covariates between IgE groups using an IPTW approach^20^, which can provide less biased and more precise estimates than multivariable regression in highly stratified analyses with few events per confounder^45^. Age, sex, and race/ethnicity have been consistently associated with measured IgE levels and glioma risk. As such, we felt it essential to balance these strong confounders between IgE groups to best assess the direct implications of IgE antibody levels on glioma risk and survival in our observational data. Taken together with known sex differences in immune responses, these results suggest that IgE-related immunologic features may contribute to both the higher risk of glioma in males and the female survival advantage observed among GBM patients. Our results provide support for further investigations including functional studies to assess the therapeutic potential of IgE-based immunotherapies for GBM.

## Author contributions

GG and SSF conceived the project. GG, TN, LK and SSF advised on the methodology. GG, LM, HMH, TR, JLW, JKW, AMM, MW and SSF were involved in primary data collection and curation. GG performed the main analyses. TN, GG, LK and SSF drafted the manuscript and all authors contributed to, reviewed and approved the final manuscript.

## Data Availability

All data from this study are a part of the University of California, San Francisco Adult Glioma Study and are available upon request from dbGap (accession phs001497.v2.p1).

https://www.ncbi.nlm.nih.gov/projects/gap/cgi-bin/study.cgi?study_id=phs001497.v2.p1

## Acknowledgements

The authors wish to acknowledge study participants, the clinicians, and the research staff at the participating medical centers, the UCSF Cancer Registry, and the UCSF Neurosurgery Tissue Bank. The collection of cancer incidence data used in this study was supported by the California Department of Public Health pursuant to California Health and Safety Code Section 103885; Centers for Disease Control and Prevention’s (CDC) National Program of Cancer Registries, under cooperative agreement 5NU58DP006344; the National Cancer Institute’s Surveillance, Epidemiology and End Results Program under contract HHSN261201800032I awarded to the University of California, San Francisco, contract HHSN261201800015I awarded to the University of Southern California, and contract HHSN261201800009I awarded to the Public Health Institute, Cancer Registry of Greater California. The ideas and opinions expressed herein are those of the author(s) and do not necessarily reflect the opinions of the State of California, Department of Public Health, the National Cancer Institute, and the Centers for Disease Control and Prevention or their Contractors and Subcontractors. All analyses, interpretations, and conclusions reached in this manuscript from the mortality data are those of the author(s) and not the State of California Department of Public Health. R01CA52689, P50CA097257, R01CA126831, R01CA139020 provided funding for the UCSF Adult Glioma Study. R01CA266676 (to SSF) provided funding for GG, HMH, and SSF during the time of study. R00CA246076 (to LK) provided funding to LK during the time of study. The funder did not play a role in the design of the study; the collection, analysis, and interpretation of the data; the writing of the manuscript; and the decision to submit the manuscript for publication.

