## Supplemental Tables and Figures for "Association of immunoglobulin E levels with glioma risk and survival"

**Contents**

- **Supplementary Table 1: Summary of demographic and treatment differences among adult glioma cases according to serum IgE measurements.**
- **Supplementary Table 2: Summary of demographic differences among adult non-glioma controls according to serum IgE measurements.**
- **Supplementary Table 3: Summary of available IgE measurements stratified by University of California, San Francisco Adult Glioma Study recruitment series.**
- **Supplementary Table 4: Associations between IgE levels and grade IV IDH-wildtype glioma risk stratified by sex.**
- **Supplementary Table 5: Heterogeneity tests of glioma risk associations between male and female participants.**
- **Supplementary Table 6: Associations between IgE levels and glioma risk stratified by sex and molecular subtype for self-reported White participants.**
- **Supplementary Table 7: Associations between IgE levels and survival among glioma cases stratified by sex and molecular subtype.**
- **Supplementary Table 8: Associations between IgE levels and survival for grade IV IDH-wildtype glioma cases.**
- **Supplementary Table 9: Heterogeneity tests of survival associations between male and female glioma cases.**
- **Supplementary Table 10: Associations between IgE levels and survival among glioma cases stratified by sex and molecular subtype for self-reported White individuals.**
- **Supplementary Table 11: Population attributable fractions (PAF) for IDH-WT survival stratified by IgE class and sex.**
- **Supplementary Figure 1: Population attributable fractions (PAF) for IDH-WT survival stratified by sex and follow-up time.**

**Supplementary Table 1: Summary of demographic and treatment differences among adult glioma cases according to serum IgE measurements.**

|  | Total IgE | | | Respiratory IgE | | | Food IgE | | |
| --- | --- | --- | --- | --- | --- | --- | --- | --- | --- |
|  | Above | Normal | P value | Positive | Negative | P value | Positive | Negative | P value |
| Mean age | 50.4 | 51.2 | 0.26 | 47.7 | 52.8 | <.001 | 52.1 | 50.7 | 0.34 |
| Sex |  |  |  |  |  |  |  |  |  |
| Male | 538 | 467 |  | 443 | 564 |  | 56 | 952 |  |
| Female | 276 | 414 | <.001 | 229 | 458 | <.001 | 26 | 660 | 0.12 |
| Race/ethnicity |  |  |  |  |  |  |  |  |  |
| White | 699 | 794 |  | 571 | 922 |  | 73 | 1420 |  |
| Other^a^ | 115 | 87 | 0.009 | 101 | 100 | 0.001 | 9 | 192 | 0.94 |
| Chemotherapy |  |  |  |  |  |  |  |  |  |
| Yes | 596 | 646 |  | 500 | 743 |  | 67 | 1176 |  |
| No | 218 | 235 | 1.00 | 172 | 279 | 0.47 | 15 | 436 | 0.10 |
| Temozolomide |  |  |  |  |  |  |  |  |  |
| Yes | 523 | 569 |  | 431 | 663 |  | 59 | 1036 |  |
| No | 291 | 312 | 0.93 | 241 | 359 | 0.80 | 23 | 576 | 0.19 |
| Radiation |  |  |  |  |  |  |  |  |  |
| Yes | 658 | 718 |  | 531 | 845 |  | 70 | 1306 |  |
| No | 156 | 163 | 0.77 | 141 | 177 | 0.07 | 12 | 306 | 0.40 |
| Dexamethasone |  |  |  |  |  |  |  |  |  |
| Yes | 302 | 349 |  | 247 | 405 |  | 33 | 619 |  |
| No | 512 | 532 | 0.31 | 425 | 617 | 0.26 | 49 | 993 | 0.83 |
| Total | 814 | 881 |  | 672 | 1022 |  | 82 | 1612 |  |

Chi-squared tests were used to assess differences between categorical variables. The Wilcoxon rank sum test was used to examine differences in the age distribution. ^a^Other includes Asians, Blacks or African Americans, Native Americans, Pacific Islanders and individuals with no reported race and ethnicity.

**Supplementary Table 2: Summary of demographic differences among adult non-glioma controls according to serum IgE measurements.**

|  | Total IgE | | | Respiratory IgE | | | Food IgE | | |
| --- | --- | --- | --- | --- | --- | --- | --- | --- | --- |
|  | Above | Normal | P value | Positive | Negative | P value | Positive | Negative | P value |
| Mean age | 56.1 | 56.4 | 0.86 | 54.0 | 57.8 | <.001 | 58.8 | 56.0 | 0.06 |
| Sex |  |  |  |  |  |  |  |  |  |
| Male | 376 | 212 |  | 267 | 321 |  | 53 | 535 |  |
| Female | 281 | 270 | <.001 | 209 | 342 | 0.01 | 37 | 514 | 0.18 |
| Race/ethnicity |  |  |  |  |  |  |  |  |  |
| White | 533 | 448 |  | 385 | 596 |  | 68 | 913 |  |
| Other^a^ | 124 | 34 | <.001 | 91 | 67 | <.001 | 22 | 136 | 0.004 |
| Total | 657 | 482 |  | 476 | 663 |  | 90 | 1049 |  |

Chi-squared tests were used to assess differences between categorical variables. The Wilcoxon rank sum test was used to examine differences in the age distribution. ^a^Other includes Asians, Blacks or African Americans, Native Americans, Pacific Islanders and individuals with no reported race and ethnicity.

**Supplementary Table 3: Summary of available IgE measurements stratified by University of California, San Francisco Adult Glioma Study recruitment series.**

|  | Total IgE | | Respiratory IgE | | Food IgE | |
| --- | --- | --- | --- | --- | --- | --- |
|  | Above | Normal | Positive | Negative | Positive | Negative |
| Series 2 (’97-’99) | 258 | 257 | 199 | 314 | 35 | 478 |
| Series 3 (’01-’04) | 673 | 668 | 514 | 828 | 86 | 1256 |
| Series 4 (’06-’10) | 540 | 438 | 435 | 543 | 51 | 927 |
| Total | 1471 | 1363 | 1148 | 1685 | 172 | 2661 |

**Supplementary Table 4: Associations between IgE levels and grade IV IDH-wildtype glioma risk stratified by sex.**

|  | Controls | Grade IV IDH-wildtype | | |
| --- | --- | --- | --- | --- |
|  | N (%) | N (%) | RR (95% CI) | P value |
| Combined |  |  |  |  |
| Total IgE |  |  |  |  |
| Normal | 482 (42) | 392 (54) | 1.00 |  |
| Above normal | 657 (58) | 335 (46) | 0.72 (0.65, 0.81) | <.001 |
| Respiratory IgE |  |  |  |  |
| Negative | 663 (58) | 478 (66) | 1.00 |  |
| Positive | 476 (42) | 249 (34) | 0.83 (0.74, 0.93) | 0.002 |
| Food IgE |  |  |  |  |
| Negative | 1049 (92) | 682 (94) | 1.00 |  |
| Positive | 90 (8) | 45 (6) | 0.87 (0.78, 0.98) | 0.02 |
| Males |  |  |  |  |
| Total IgE |  |  |  |  |
| Normal | 212 (36) | 212 (47) | 1.00 |  |
| Above normal | 376 (64) | 237 (53) | 0.74 (0.65, 0.85) | <.001 |
| Respiratory IgE |  |  |  |  |
| Negative | 321 (55) | 285 (62) | 1.00 |  |
| Positive | 267 (45) | 177 (38) | 0.88 (0.77, 1.01) | 0.07 |
| Food IgE |  |  |  |  |
| Negative | 535 (91) | 434 (94) | 1.00 |  |
| Positive | 53 (9) | 29 (6) | 0.81 (0.70, 0.93) | 0.003 |
| Females |  |  |  |  |
| Total IgE |  |  |  |  |
| Normal | 270 (49) | 167 (63) | 1.00 |  |
| Above normal | 281 (51) | 98 (37) | 0.69 (0.56, 0.84) | <.001 |
| Respiratory IgE |  |  |  |  |
| Negative | 342 (62) | 193 (73) | 1.00 |  |
| Positive | 209 (38) | 72 (27) | 0.74 (0.60, 0.91) | 0.005 |
| Food IgE |  |  |  |  |
| Negative | 514 (93) | 248 (94) | 1.00 |  |
| Positive | 37 (7) | 16 (6) | 0.98 (0.80, 1.19) | 0.82 |

Risk ratios (RR) and 95% confidence intervals (CI) were estimated with weighted logistic regression models using inverse probability weighting for age, sex (included in combined analysis only) and self-reported race/ethnicity. Models are also adjusted for AGS recruitment series.

**Supplementary Table 5: Heterogeneity tests of glioma risk associations between male and female participants.**

|  | IDH-wildtype | | | IDH-mutant | | | Grade IV IDH-wildtype | | |
| --- | --- | --- | --- | --- | --- | --- | --- | --- | --- |
|  | I^2^ | Q | P value | I^2^ | Q | P value | I^2^ | Q | P value |
| Total IgE | 0% | 0.31 | 0.58 | 0% | 0.62 | 0.43 | 0% | 0.40 | 0.54 |
| Respiratory IgE | 0% | 0.65 | 0.42 | 60.2% | 2.51 | 0.11 | 47.5% | 1.90 | 0.17 |
| Food IgE | 73.4% | 3.75 | 0.05 | — | — | — | 57.5% | 2.35 | 0.13 |

Heterogeneity tests were performed for glioma risk associations between male and female participants. I^2^ index and Cochran’s Q statistic are reported. Results are not reported for IDH-mutant glioma and food IgE due to insufficient female participants with positive food IgE.

**Supplementary Table 6: Associations between IgE levels and glioma risk stratified by sex and molecular subtype for self-reported White participants.**

|  | **Controls** | **IDH-wildtype** | | | **IDH-mutant** | | |
| --- | --- | --- | --- | --- | --- | --- | --- |
|  | **N (%)** | **N (%)** | **RR (95% CI)** | **P value** | **N (%)** | **RR (95% CI)** | **P value** |
| Combined |  |  |  |  |  |  |  |
| Total IgE |  |  |  |  |  |  |  |
| Normal | 448 (45) | 435 (53) | 1.00 |  | 182 (53) | 1.00 |  |
| Above normal | 533 (55) | 391 (47) | 0.83 (0.76, 0.92) | <.001 | 159 (47) | 0.87 (0.74, 1.02) | 0.08 |
| Respiratory IgE |  |  |  |  |  |  |  |
| Negative | 596 (61) | 532 (64) | 1.00 |  | 199 (58) | 1.00 |  |
| Positive | 385 (39) | 294 (36) | 0.89 (0.81, 0.99) | 0.03 | 142 (42) | 0.94 (0.80, 1.09) | 0.39 |
| Food IgE |  |  |  |  |  |  |  |
| Negative | 913 (93) | 777 (94) | 1.00 |  | 331 (97) | 1.00 |  |
| Positive | 68 (7) | 49 (6) | 0.92 (0.84, 1.02) | 0.12 | 10 (3) | 0.57 (0.47, 0.70) | <.001 |
| Males |  |  |  |  |  |  |  |
| Total IgE |  |  |  |  |  |  |  |
| Normal | 201 (39) | 244 (47) | 1.00 |  | 94 (47) | 1.00 |  |
| Above normal | 320 (61) | 271 (53) | 0.84 (0.74, 0.94) | 0.004 | 106 (53) | 0.90 (0.73, 1.10) | 0.29 |
| Respiratory IgE |  |  |  |  |  |  |  |
| Negative | 290 (56) | 312 (61) | 1.00 |  | 107 (53) | 1.00 |  |
| Positive | 231 (44) | 203 (39) | 0.91 (0.80, 1.03) | 0.12 | 94 (47) | 0.96 (0.80, 1.16) | 0.66 |
| Food IgE |  |  |  |  |  |  |  |
| Negative | 478 (92) | 485 (94) | 1.00 |  | 192 (96) | 1.00 |  |
| Positive | 43 (8) | 31 (6) | 0.84 (0.74, 0.96) | 0.01 | 9 (4) | 0.68 (0.54, 0.86) | 0.002 |
| Females |  |  |  |  |  |  |  |
| Total IgE |  |  |  |  |  |  |  |
| Normal | 247 (54) | 191 (61) | 1.00 |  | 88 (62) | 1.00 |  |
| Above normal | 213 (46) | 120 (39) | 0.83 (0.69, 0.98) | 0.03 | 53 (38) | 0.83 (0.63, 1.08) | 0.17 |
| Respiratory IgE |  |  |  |  |  |  |  |
| Negative | 306 (67) | 220 (71) | 1.00 |  | 92 (66) | 1.00 |  |
| Positive | 154 (33) | 91 (29) | 0.87 (0.73, 1.04) | 0.12 | 48 (34) | 0.90 (0.68, 1.18) | 0.04 |
| Food IgE |  |  |  |  |  |  |  |
| Negative | 435 (95) | 292 (94) | 1.00 |  | 139 (100) | — |  |
| Positive | 25 (5) | 18 (6) | 1.06 (0.90, 1.25) | 0.46 | 1 (0) | — | — |

Risk ratios (RR) and 95% confidence intervals (CI) were estimated with weighted logistic regression models using inverse probability weighting for age, and sex (included in combined analysis only). Models are also adjusted for AGS recruitment series. Risk analysis for food IgE among female patients with IDH-mutant glioma is not reported due to insufficient individuals with positive food IgE.

**Supplementary Table 7: Associations between IgE levels and survival among glioma cases stratified by sex and molecular subtype.**

|  | IDH-wildtype | | | | IDH-mutant | | | |
| --- | --- | --- | --- | --- | --- | --- | --- | --- |
|  | Cases (Deaths) | Median Survival  (Months) | RR (95% CI) | P value | Cases (Deaths) | Median Survival  (Months) | RR (95% CI) | P value |
| Combined |  |  |  |  |  |  |  |  |
| Total IgE |  |  |  |  |  |  |  |  |
| Normal | 400 (382) | 15.6 | 1.00 |  | 155 (96) | 133 | 1.00 |  |
| Above normal | 362 (341) | 17.1 | 0.84 (0.71, 0.98) | 0.03 | 145 (80) | 160 | 0.89 (0.63, 1.26) | 0.50 |
| Respiratory IgE |  |  |  |  |  |  |  |  |
| Negative | 491 (472) | 15.4 | 1.00 |  | 165 (98) | 136 | 1.00 |  |
| Positive | 271 (251) | 17.6 | 0.79 (0.67, 0.93) | 0.005 | 135 (78) | 142 | 0.92 (0.66, 1.29) | 0.63 |
| Food IgE |  |  |  |  |  |  |  |  |
| Negative | 718 (680) | 16.2 | 1.00 |  | 292 (172) | 138 | — |  |
| Positive | 44 (43) | 19.2 | 0.99 (0.75, 1.31) | 0.95 | 8 (5) | 162 | — | — |
| Male |  |  |  |  |  |  |  |  |
| Total IgE |  |  |  |  |  |  |  |  |
| Normal | 220 (215) | 14.8 | 1.00 |  | 83 (54) | 127 | 1.00 |  |
| Above normal | 256 (242) | 16.0 | 0.86 (0.71, 1.04) | 0.11 | 97 (55) | 147 | 0.86 (0.54, 1.37) | 0.53 |
| Respiratory IgE |  |  |  |  |  |  |  |  |
| Negative | 286 (278) | 14.5 | 1.00 |  | 89 (59) | 116 | 1.00 |  |
| Positive | 190 (179) | 16.5 | 0.83 (0.67, 1.03) | 0.09 | 92 (51) | 160 | 0.76 (0.49, 1.18) | 0.22 |
| Food IgE |  |  |  |  |  |  |  |  |
| Negative | 450 (431) | 15.8 | 1.00 |  | 173 (105) | 136 | 1.00 |  |
| Positive | 27 (27) | 14.7 | 1.24 (0.87, 1.78) | 0.23 | 8 (5) | 162 | 1.58 (0.81, 3.11) | 0.18 |
| Female |  |  |  |  |  |  |  |  |
| Total IgE |  |  |  |  |  |  |  |  |
| Normal | 184 (169) | 16.3 | 1.00 |  | 72 (42) | 137 | 1.00 |  |
| Above normal | 111 (102) | 22.5 | 0.76 (0.59, 0.98) | 0.04 | 48 (25) | 174 | 0.77 (0.44, 1.34) | 0.36 |
| Respiratory IgE |  |  |  |  |  |  |  |  |
| Negative | 209 (196) | 16.4 | 1.00 |  | 76 (40) | 174 | 1.00 |  |
| Positive | 86 (75) | 23.3 | 0.75 (0.57, 0.98) | 0.04 | 43 (27) | 137 | 1.57 (0.92, 2.68) | 0.10 |
| Food IgE |  |  |  |  |  |  |  |  |
| Negative | 277 (254) | 17.7 | 1.00 |  | 119 (67) | — | — |  |
| Positive | 17 (16) | 20.7 | 0.95 (0.62, 1.46) | 0.81 | 0 (0) | — | — | — |

Risk ratios (RR) and 95% confidence intervals (CI) were estimated with weighted Cox regression models using inverse probability weighting for age, self-reported race/ethnicity and chemotherapy use. Model also include sex (included in combined analysis only), dexamethasone use, radiation use, surgery type (resection/biopsy), 1p19q codeletion, and *TERT* mutation, tumor grade and AGS recruitment series as covariates. Survival analyses for food IgE and IDH-mutant glioma (combined and female only) are not reported due to insufficient individuals with positive food IgE.

**Supplementary Table 8: Associations between IgE levels and survival for grade IV IDH-wildtype glioma cases.**

|  | Grade IV IDH-wildtype | | | |
| --- | --- | --- | --- | --- |
|  | Cases (Deaths) | Median Survival (Months) | RR (95% CI) | P value |
| Combined |  |  |  |  |
| Total IgE |  |  |  |  |
| Normal | 355 (347) | 15.2 | 1.00 |  |
| Above normal | 311 (303) | 16.0 | 0.83 (0.71, 0.98) | 0.03 |
| Respiratory IgE |  |  |  |  |
| Negative | 435 (426) | 14.7 | 1.00 |  |
| Positive | 231 (224) | 16.7 | 0.80 (0.68, 0.95) | 0.01 |
| Food IgE |  |  |  |  |
| Negative | 624 (609) | 15.6 | 1.00 |  |
| Positive | 42 (41) | 17.5 | 0.97 (0.72, 1.30) | 0.83 |
| Male |  |  |  |  |
| Total IgE |  |  |  |  |
| Normal | 198 (196) | 14.3 | 1.00 |  |
| Above normal | 219 (214) | 14.7 | 0.86 (0.71, 1.05) | 0.14 |
| Respiratory IgE |  |  |  |  |
| Negative | 255 (251) | 13.6 | 1.00 |  |
| Positive | 162 (159) | 15.8 | 0.84 (0.68, 1.05) | 0.13 |
| Food IgE |  |  |  |  |
| Negative | 392 (285) | 14.6 | 1.00 |  |
| Positive | 26 (26) | 14.2 | 1.25 (0.88, 1.81) | 0.21 |
| Female |  |  |  |  |
| Total IgE |  |  |  |  |
| Normal | 157 (151) | 15.6 | 1.00 |  |
| Above normal | 92 (89) | 21.0 | 0.73 (0.55, 0.97) | 0.03 |
| Respiratory IgE |  |  |  |  |
| Negative | 180 (175) | 15.6 | 1.00 |  |
| Positive | 69 (65) | 21.0 | 0.77 (0.57, 1.04) | 0.09 |
| Food IgE |  |  |  |  |
| Negative | 232 (224) | 16.9 | 1.00 |  |
| Positive | 16 (15) | 21.7 | 0.83 (0.51, 1.37) | 0.46 |

Risk ratios (RR) and 95% confidence intervals (CI) were estimated with weighted Cox regression models using inverse probability weighting for age, self-reported race/ethnicity and chemotherapy use. Model also include sex (included in combined analysis only), dexamethasone use, radiation use, surgery type (resection/biopsy), 1p19q codeletion, and *TERT* mutation, and AGS recruitment series as covariates.

**Supplementary Table 9: Heterogeneity tests of survival associations between male and female glioma cases.**

|  | IDH-wildtype | | | IDH-mutant | | | Grade IV IDH-wildtype | | |
| --- | --- | --- | --- | --- | --- | --- | --- | --- | --- |
|  | I^2^ | Q | P value | I^2^ | Q | P value | I^2^ | Q | P value |
| Total IgE | 0% | 0.47 | 0.47 | 0% | 0.09 | 0.77 | 0% | 0.86 | 0.35 |
| Respiratory IgE | 0% | 0.53 | 0.53 | 75.8% | 4.14 | 0.04 | 0% | 0.25 | 0.61 |
| Food IgE | 0% | 0.90 | 0.34 | — | — | — | 43.3% | 1.76 | 0.18 |

Heterogeneity tests were performed for survival associations between male and female glioma cases. I^2^ index and Cochran’s Q statistic are reported. Results are not reported for IDH-mutant glioma and food IgE due to insufficient female cases with positive food IgE.

**Supplementary Table 10: Associations between IgE levels and survival among glioma cases stratified by sex and molecular subtype for self-reported White individuals.**

|  | **IDH-wildtype** | | | | **IDH-mutant** | | | |
| --- | --- | --- | --- | --- | --- | --- | --- | --- |
|  | **Cases (Deaths)** | **Median Survival**  **(Months)** | **RR (95% CI)** | **P value** | **Cases (Deaths)** | **Median Survival**  **(Months)** | **RR (95% CI)** | **P value** |
| Combined |  |  |  |  |  |  |  |  |
| Total IgE |  |  |  |  |  |  |  |  |
| Normal | 368 (350) | 15.7 | 1.00 |  | 139 (87) | 130 | 1.00 |  |
| Above normal | 320 (302) | 16.8 | 0.88 (0.75, 1.04) | 0.14 | 125 (74) | 142 | 0.95 (0.66, 1.37) | 0.80 |
| Respiratory IgE |  |  |  |  |  |  |  |  |
| Negative | 450 (431) | 15.4 | 1.00 |  | 148 (91) | 126 | 1.00 |  |
| Positive | 238 (221) | 17.4 | 0.83 (0.70, 0.99) | 0.04 | 116 (71) | 137 | 0.97 (0.67, 1.39) | 0.85 |
| Food IgE |  |  |  |  |  |  |  |  |
| Negative | 648 (612) | 16.2 | 1.00 |  | 257 (157) | 133 | — |  |
| Positive | 40 (40) | 19.2 | 1.00 (0.76, 1.33) | 0.99 | 7 (5) | 161 | — | — |
| Male |  |  |  |  |  |  |  |  |
| Total IgE |  |  |  |  |  |  |  |  |
| Normal | 203 (198) | 14.9 | 1.00 |  | 76 (49) | 127 | 1.00 |  |
| Above normal | 229 (216) | 15.8 | 0.87 (0.71, 1.07) | 0.18 | 83 (51) | 137 | 1.03 (0.65, 1.65) | 0.89 |
| Respiratory IgE |  |  |  |  |  |  |  |  |
| Negative | 263 (255) | 14.5 | 1.00 |  | 81 (53) | 116 | 1.00 |  |
| Positive | 169 (159) | 16.5 | 0.86 (0.70, 1.07) | 0.18 | 79 (48) | 137 | 0.84 (0.54, 1.33) | 0.47 |
| Food IgE |  |  |  |  |  |  |  |  |
| Negative | 407 (389) | 15.8 | 1.00 |  | 153 (96) | 126 | 1.00 |  |
| Positive | 26 (26) | 14.9 | 1.21 (0.84, 1.76) | 0.30 | 7 (5) | 161 | 1.66 (0.87, 3.19) | 0.13 |
| Female |  |  |  |  |  |  |  |  |
| Total IgE |  |  |  |  |  |  |  |  |
| Normal | 166 (153) | 16.0 | 1.00 |  | 63 (38) | 137 | 1.00 |  |
| Above normal | 91 (86) | 21.3 | 0.85 (0.65, 1.13) | 0.26 | 42 (23) | 146 | 0.79 (0.44, 1.43) | 0.44 |
| Respiratory IgE |  |  |  |  |  |  |  |  |
| Negative | 187 (176) | 16.0 | 1.00 |  | 67 (38) | 163 | 1.00 |  |
| Positive | 70 (63) | 21.5 | 0.83 (0.62, 1.11) | 0.20 | 37 (23) | 142 | 1.32 (0.75, 2.35) | 0.33 |
| Food IgE |  |  |  |  |  |  |  |  |
| Negative | 242 (224) | 17.2 | 1.00 |  | 104 (61) | — | — |  |
| Positive | 14 (14) | 21.7 | 0.90 (0.58, 1.39) | 0.63 | 0 (0) | — | — | — |

Risk ratios (RR) and 95% confidence intervals (CI) were estimated with weighted Cox regression models using inverse probability weighting for age and chemotherapy use. Model also include sex (included in combined analysis only), dexamethasone use, radiation use, surgery type (resection/biopsy), 1p19q codeletion, and *TERT* mutation, tumor grade and AGS recruitment series as covariates. Survival analyses for food IgE and IDH-mutant glioma (combined and female only) are not reported due to insufficient individuals with positive food IgE.

**Supplementary Table 11: Population attributable fractions (PAF) for IDH-WT survival stratified by IgE class and sex.**

|  | Total IgE | | Respiratory IgE | |
| --- | --- | --- | --- | --- |
|  | AF (95% CI) | P value | AF (95% CI) | P value |
| Combined |  |  |  |  |
| 6 months |  |  |  |  |
| Age at diagnosis | 0.26 (0.19, 0.33) | <.001 | 0.25 (0.18, 0.33) | <.001 |
| Surgery | 0.08 (0.02, 0.13) | 0.01 | 0.08 (0.03, 0.13) | 0.003 |
| Chemotherapy | 0.08 (0.01, 0.15) | 0.02 | 0.08 (0.01, 0.14) | 0.02 |
| IgE | 0.08 (0.00, 0.16) | 0.04 | 0.14 (0.04, 0.23) | 0.004 |
| 12 months |  |  |  |  |
| Age at diagnosis | 0.21 (0.15, 0.27) | <.001 | 0.20 (0.14, 0.26) | <.001 |
| Surgery | 0.05 (0.02, 0.09) | 0.005 | 0.05 (0.02, 0.08) | 0.001 |
| Chemotherapy | 0.05 (0.01, 0.10) | 0.02 | 0.05 (0.01, 0.10) | 0.03 |
| IgE | 0.06 (0.00, 0.13) | 0.04 | 0.11 (0.03, 0.18) | 0.005 |
| Male |  |  |  |  |
| 6 months |  |  |  |  |
| Age at diagnosis | 0.24 (0.15, 0.34) | <.001 | 0.24 (0.15, 0.33) | <.001 |
| Surgery | 0.10 (0.01, 0.19) | 0.03 | 0.10 (0.02, 0.18) | 0.02 |
| Chemotherapy | 0.10 (0.03, 0.16) | 0.003 | 0.09 (0.03, 0.16) | 0.005 |
| IgE | 0.06 (-0.03, 0.14) | 0.17 | 0.11 (-0.01, 0.23) | 0.08 |
| 12 months |  |  |  |  |
| Age at diagnosis | 0.18 (0.11, 0.26) | <.001 | 0.18 (0.11, 0.26) | <.001 |
| Surgery | 0.06 (0.01, 0.10) | 0.02 | 0.06 (0.01, 0.10) | 0.009 |
| Chemotherapy | 0.06 (0.02, 0.10) | 0.004 | 0.06 (0.02, 0.10) | 0.005 |
| IgE | 0.04 (-0.02, 0.11) | 0.17 | 0.08 (-0.01, 0.18) | 0.09 |
| Female |  |  |  |  |
| 6 months |  |  |  |  |
| Age at diagnosis | 0.34 (0.22, 0.45) | <.001 | 0.33 (0.21, 0.44) | <.001 |
| Surgery | 0.07 (0.01, 0.12) | 0.02 | 0.07 (0.01, 0.12) | 0.02 |
| Chemotherapy | 0.07 (-0.06, 0.20) | 0.30 | 0.06 (-0.07, 0.19) | 0.36 |
| IgE | 0.15 (0.02, 0.28) | 0.03 | 0.19 (0.04, 0.34) | 0.01 |
| 12 months |  |  |  |  |
| Age at diagnosis | 0.29 (0.19, 0.39) | <.001 | 0.28 (0.17, 0.38) | <.001 |
| Surgery | 0.05 (0.01, 0.09) | 0.01 | 0.05 (0.01, 0.09) | 0.02 |
| Chemotherapy | 0.05 (-0.05, 0.14) | 0.31 | 0.04 (-0.05, 0.14) | 0.37 |
| IgE | 0.13 (0.01, 0.24) | 0.03 | 0.16 (0.03, 0.29) | 0.01 |

PAF estimates for age at diagnosis (≥58 years/<58 years), surgery (biopsy/resection), chemotherapy use (yes/no), and IgE (total IgE: above normal/normal; respiratory IgE: positive/negative) were derived from Cox proportional hazard regression models adjusted for sex (combined only), self-reported race and ethnicity, radiation use, 1p19q codeletion status, *TERT* mutation status, and strata for tumor grade. PAF estimates for radiation use are not reported for females due to insufficient individuals who received radiation therapy. PAF estimated are reported for 6 months and 12 months of follow-up time.

**Supplementary Figure 1: Population attributable fractions (PAF) for IDH-WT survival stratified by sex and follow-up time.** PAF estimates for age at diagnosis (<58 years/≥58 years), surgery (resection/biopsy), chemotherapy use (yes/no), and total IgE (above normal/normal) were derived from Cox proportional hazard regression models adjusted for sex (combined only), self-reported race and ethnicity, radiation use, 1p19q codeletion status, *TERT* mutation status, and strata for tumor grade. PAF estimates are reported for 6 months and 12 months of follow-up time. Estimates that do not achieve P<0.05 are indicated by a striped pattern.

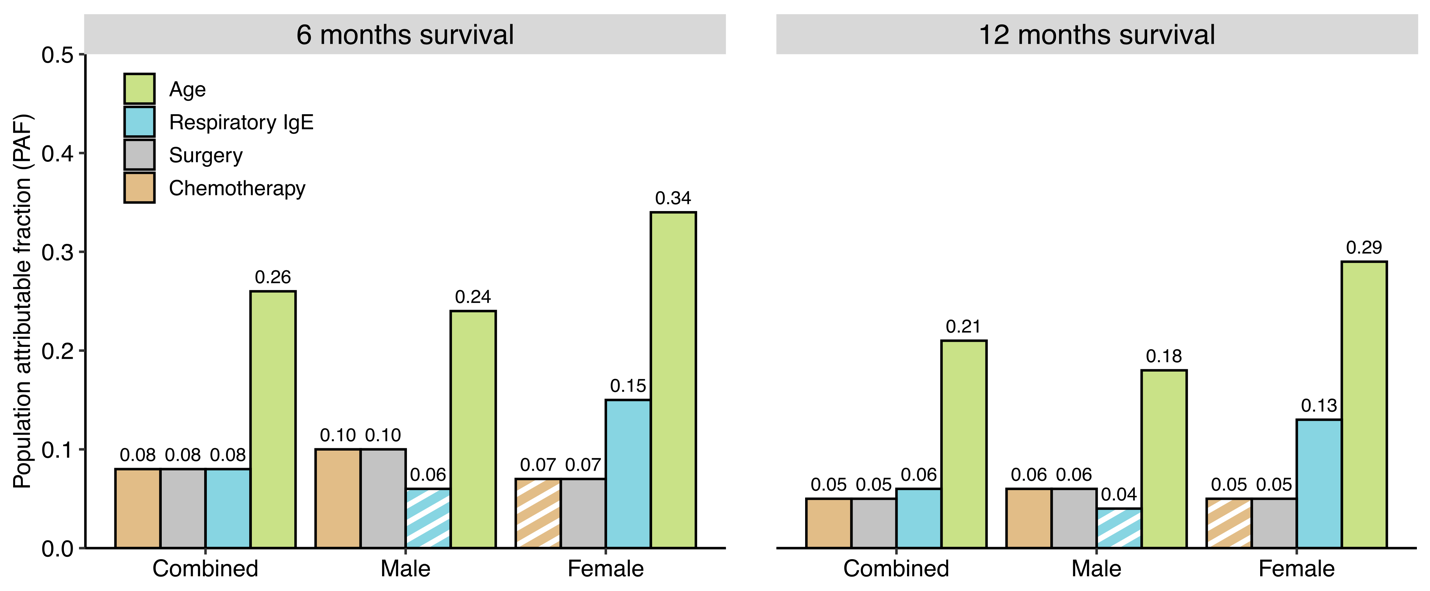
